# A UK national survey of prophylactic platelet transfusion thresholds in non-bleeding, critically ill adults

**DOI:** 10.1101/2020.09.02.20186700

**Authors:** Akshay Shah, Doug W Gould, James Doidge, Paul R Mouncey, David A Harrison, J Duncan Young, Simon J Stanworth, Peter J Watkinson, on behalf of the Threshold for Platelets (T4P) Investigators

**Affiliations:** Radcliffe Department of Medicine, University of Oxford, Oxford, UK; Adult Intensive Care Unit, Oxford University Hospitals NHS Foundation Trust, Oxford, UK; Clinical Trials Unit, Intensive Care National Audit & Research Centre (ICNARC), London, UK; Nuffield Department of Clinical Neurosciences, University of Oxford, Oxford, UK; NHS Blood & Transplant, Oxford, UK

## Abstract

Thrombocytopaenia is common in critically ill patients and associated with poor clinical outcomes. Current guideline recommendations for prophylactic platelet transfusions, to prevent bleeding in critically ill patients with thrombocytopaenia, are based on observational data. Recent studies conducted in non-critically ill patients have demonstrated harm associated with platelet transfusions and have also called into question the efficacy of platelet transfusion. To date, there are no well-conducted randomised controlled trials (RCTs) evaluating platelet transfusion in critically ill patients. To inform the design of such an RCT, we sought to characterise current clinical practice across four commonly encountered scenarios in non-bleeding critically ill adult patients with thrombocytopaenia. An online survey link was sent to Clinical Directors and contacts of all adult general ICUs participating in the Intensive Care National Audit and Research Centre Case Mix Programme national clinical audit (n=200). The survey collected data regarding the respondents place of work, training grade and their current individual practice and possible limits of equipoise for prescribing prophylactic platelet transfusions across four scenarios: prophylaxis but with no procedure planned (NPP); ultrasound guided insertion of a right internal jugular central venous catheter (JVI); percutaneous tracheostomy (PT); and surgery with a low bleeding risk (SLBR). After excluding nine responses with missing data on all four of the main questions, responses were received from 99 staff, covering 78 ICUs (39.0% of 200 ICUs invited to participate). While nearly all respondents (98.0%) indicated a platelet transfusion threshold of 30 x 10^9/L or less for patients with no planned procedure, thresholds for planned procedures varied widely and centred at medians of 40 x 10^9/L for JVI (range: 10 to 70), 50 x 10^9/L for SLBR (range: 10 to 100)  and 70 x 10^9/L for PT (range: 20 to greater than 100). Current platelet transfusion practice in UK ICUs prior to invasive procedures with relatively low bleeding risks is highly variable. Well-designed studies are needed to determine the optimal platelet transfusion thresholds in critical care.

---

Dear editor

Thrombocytopaenia is well recognised in intensive care unit (ICU) patients. Studies show 8.3% to 67.6% of patients admitted to ICU have platelet counts below 50 or 100 × 10^9^/L^1^ The wide variation in the reported prevalence of thrombocytopaenia likely reflects the broad case mix, timing of measurement and lack of a standardised definition. Despite this, thrombocytopaenia is associated with an increased risk of death, longer ICU and hospital length of stays, and higher requirements for organ support.^1^

Platelet transfusions are prescribed prophylactically to ICU patients with thrombocytopaenia to mitigate the potential risk of spontaneous bleeding and/or bleeding prior to an invasive procedure. ICU services are the second biggest consumer of platelet transfusions in the United Kingdom (UK), behind cancer services, but there are no well conducted randomised controlled trials (RCTs) in ICU patients to inform this practice.^2^ Data from observational studies in ICU patients suggests that prophylactic platelet transfusions do not reduce the rate of major bleeding when compared with no platelets^3^ A platelet count increment < 5 × 10^9^/L occurs following one in five platelet transfusions given to ICU patients (classified as “ineffectual”).^4^ This lack of benefit must be considered alongside the increasing recognition of harms of platelet transfusions such as increased risk of developing transfusion-associated lung injury and nosocomial infection.^3^ A recent randomised trial of prophylactic platelet transfusions in critically ill neonates demonstrated increased mortality and bleeding complications in neonates allocated to more liberal transfusion strategies.^5^

The lack of high-quality data to guide decision making has also resulted in widespread variation in practice. A large epidemiological study of 163,719 platelet transfusion episodes across twelve US hospitals found that approximately 35% of transfusion episodes occurred at counts greater than 50 × 10^9^/L,^6^ incongruous with guideline recommendations.^2,7^ Recent data from three UK ICUs also show that platelet transfusions are given at a higher median platelet count than suggested by guidelines.^8^ Therefore, appropriate use of platelet transfusions is now recognised as a research priority in critical care and well-designed RCTs are needed.^7^ To inform the design of such an RCT, we sought to characterise current clinical practice across four commonly encountered scenarios in non-bleeding critically ill adult patients with thrombocytopaenia.

## Methods

A panel of intensivists, haematologists, statisticians and trial methodologists developed and piloted an online survey. The survey collected data regarding the respondents’ place of work, training grade and their current individual practice and possible limits of equipoise for prescribing prophylactic platelet transfusions across four scenarios: prophylaxis but with no procedure planned (NPP); ultrasound guided insertion of a right internal jugular central venous catheter (JVI); percutaneous tracheostomy (PT); and surgery with a low bleeding risk (SLBR). For each scenario, respondents were given a range of ten possible responses (from 10 to ≥ 100×10^9^/L), or no response/not applicable. No personal, sensitive or confidential information about the respondents was collected.

Formal ethical approval was not required for this survey according to the Health Research Authority decision tool and guidance from the UK Data Service. Respondents were informed at the start of the survey that results could be published in aggregate.

The survey was hosted by the Survey Monkey website (http://www.survevmonkey.com). An email inviting participation was sent to Clinical Directors and contacts of all adult general ICUs participating in the Intensive Care National Audit and Research Centre Case Mix Programme national clinical audit (n = 200). All responses were anonymous. The survey window was open for two weeks. Due to the emerging Coronavirus Disease-19 (COVID-19) pandemic in the UK at the time of survey administration, no reminder or follow-up emails were sent. Descriptive summary statistical data are presented, with available case analysis of missing data.

## Results

After excluding nine responses with missing data on all four of the main questions, responses were received from 99 staff, covering 78 ICUs (39.0% of 200 ICUs invited to participate). Respondents included 40 (40.4%) clinical lead/directors, 51 (51.5%) consultants, 4 (4.0%) registrars and 4 (4.0%) other positions. The 99 respondents provided item responses for usual practice with respect to NPP (n = 99 (100.0%)), JVI (n = 91 (91.9%)), PT (n = 86 (86.9%)) and SLBR (n = 85 (85.9%)).

Responses are illustrated in **Figure 1**. While nearly all respondents (98.0%) indicated a platelet transfusion threshold of 30×10^9^/L or less for patients with no planned procedure, thresholds for planned procedures varied widely and centred at medians of 40×10^9^/L for JVI (range: 10 to 70), 50×10^9^/L for SLBR (range: 10 to ≥ 100) and 70×10^9^/L for PT (range: 20 to ≥ 100).

**Figure 1.**
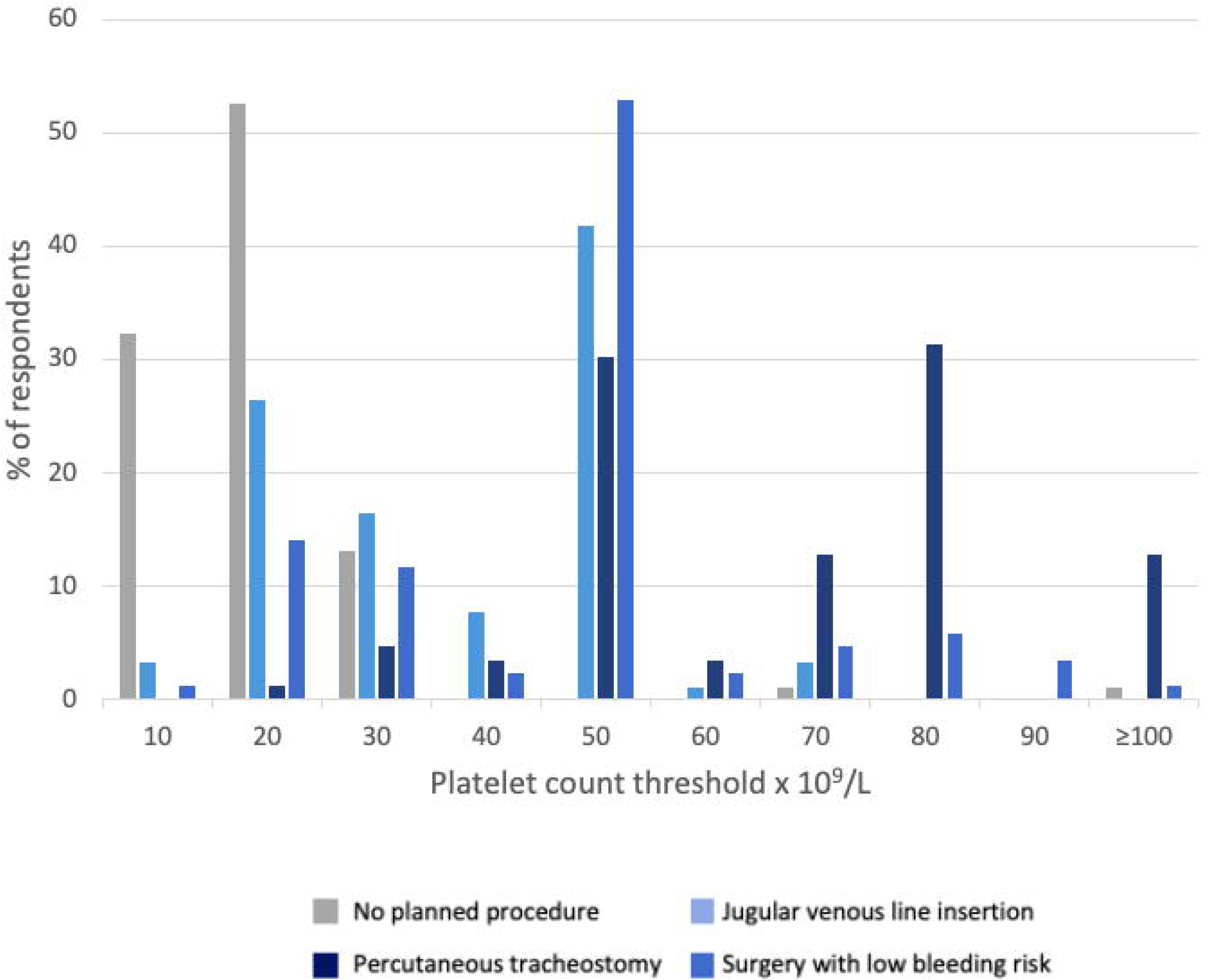
Prophylactic platelet transfusion thresholds without any planned procedures and prior to different procedures in UK ICUs.

## Discussion

The main finding of this survey is that current platelet transfusion practice in UK ICUs prior to invasive procedures with relatively low bleeding risks is highly variable. This practice variation is also apparent in the prophylaxis setting of no planned procedures. Our findings are consistent with those reported from other national^9^ and international surveys.^7^ Our findings raise questions about the role of current guidelines to standardise safe practice, which in turn are based on limited robust primary data.

The decision to transfuse platelets is influenced by patient, procedural and institutional factors. Patient factors include the presence of thrombocytopaenia and/or elevated prothrombin time, but it is now well recognised that these are poor predictors of bleeding.^10^ Procedural factors include anatomical site and use of adjuncts such as ultrasound. A meta-analysis of largely observational studies showed that bleeding complications prior to JVI placements, even in patients with thrombocytopaenia, are rare.^10^ Combined with the widespread use of ultrasound, the use of platelets for this procedure is questionable but definitive evidence is needed. Beyond haemostasis, platelets are also implicated in multiple immunological and inflammatory processes and observational studies have shown that platelet transfusions are associated with increased rates of nosocomial infection, acute respiratory distress syndrome and arterial and venous thromboses.^2,5,7^ However, causation is difficult to determine as residual confounding may exist.

The main strength of our study is the coverage of a large number of UK ICUs (n=78). Limitations include a relatively low overall response rate of approximately 40%, although reminders to complete the survey were not felt appropriate due to the impact of the COVID-19 pandemic. It is unlikely that the reported thresholds would be significantly difference with a higher response rate.

In conclusion, current UK practice of prophylactic platelet transfusion in non-bleeding ICU patients with thrombocytopaenia varies widely. Well-designed studies are needed to determine the optimal platelet transfusion thresholds in critical care, given uncertain benefits of platelets to reduce bleeding risk and concerns about harm as demonstrated in trial settings of neonatal medicine and spontaneous intracerebral haemorrhage.

## Data Availability

The dataset used and analysed for this study are available from the corresponding author on reasonable request.

## Collaborating authors

**On behalf of the Threshold for Platelets (T4P) Investigators:**

- Matteo Quartagno, MRC Clinical Trials Unit, University College London, London, UK
- Joanna Calder, Patient Representative
- Timothy S Walsh, Department of Anaesthesia, Critical Care and Pain Medicine, Royal Infirmary Edinburgh, Edinburgh, UK; Edinburgh Critical Care Research Group, University of Edinburgh, Edinburgh, UK.
- Richard Grieve, Department of Health Services, Research and Policy, London School of Hygiene and Tropical Medicine, London, UK
- Alexina Mason, Centre for Statistical Methodology, London School of Hygiene and Tropical Medicine, London, UK

## Acknowledgements

None

